# Analysis of Y Chromosome Haplogroups in Parkinson’s Disease

**DOI:** 10.1101/2022.02.28.22271633

**Authors:** Francis P. Grenn, Mary B Makarious, Sara Bandres-Ciga, Hirotaka Iwaki, Andrew Singleton, Mike Nalls, Cornelis Blauwendraat, The International Parkinson Disease Genomics Consortium

## Abstract

Parkinson’s disease is a complex neurodegenerative disorder that is about 1.5 times more prevalent in males than females. Extensive work has been done to identify the genetic risk factors behind Parkinson’s disease on autosomes and more recently on chromosome X, but work remains to be done on the male specific Y chromosome. In an effort to explore the role of the Y chromosome in Parkinson’s disease we analyzed whole genome sequencing data from the Accelerating Medicines Partnership - Parkinson’s disease initiative (1,466 cases and 1,664 controls), genotype data from NeuroX (3,491 cases and 3,232 controls) and genotype data from UKBiobank (182,517 controls, 1,892 cases, and 3,783 proxy cases) all consisting of male European ancestry samples. We classified sample Y chromosomes by haplogroup using three different tools for comparison (Snappy, Yhaplo, Y-LineageTracker), and meta-analyzed this data to identify haplogroups associated with Parkinson’s disease. This was followed up with a Y chromosome association study to identify specific variants associated with disease. We also analyzed blood based RNASeq data obtained from the Accelerating Medicines Partnership - Parkinson’s disease initiative (1,020 samples) and RNASeq data obtained from the North American Brain Expression Consortium (171 samples) to identify Y chromosome genes differentially expressed in cases, controls, specific haplogroups, and specific tissues. RNASeq analyses suggest Y chromosome gene expression differs between brain and blood tissues but does not differ significantly in cases, controls or specific haplogroups. Overall, we did not find any strong associations between Y chromosome genetics and Parkinson’s disease, suggesting the explanation for increased prevalence in males may lie elsewhere.

## 1. Introduction

Parkinson’s disease (PD) is a progressive movement disorder which includes symptoms such as tremor, slowed speech, bradykinesia, loss of balance and muscle stiffness. While age is the primary risk factor for PD, genetic and likely environmental factors contribute to PD onset and progression (Blauwendraat, Nalls, and Singleton 2020). Recent genetic studies have identified 92 PD risk variants in European and Asian ancestry populations, but common variation findings from genome wide association studies (GWAS) only account for 16-36% of heritable risk (Nalls et al. 2019; Foo et al. 2020). Environmental factors such as pesticide exposure, smoking, and caffeine intake have been linked to PD etiology, but the underlying mechanisms are not fully understood (Nandipati and Litvan 2016; Gallo et al. 2019; Ascherio et al. 2004; Noyce et al. 2012).

PD is ~1.5 times more prevalent in males than in females with European ancestry (Moisan et al. 2016). However, it is unclear whether this difference is due to environmental or genetic factors, or a combination of the two. Studies have shown that there are sex specific differences in PD clinical presentation and progression (Iwaki, Blauwendraat, et al. 2021). To date, no significant autosomal genetic differences have been found between male and female PD cases (Blauwendraat et al. 2021). A recent chromosome X GWAS did identify potential GWAS hits but no significant differences in PD risk between sexes (Le Guen et al. 2021). This leaves the male specific chromosome Y as a potential candidate for sex specific PD risk.

Chromosome Y is frequently excluded from large scale genome wide association studies due to its size and unique characteristics (Parker, Mesut Erzurumluoglu, and Rodriguez 2020). The Y chromosome makes up ~2% of the total DNA in a human male cell. Additionally, quality control filters typically used in GWAS, such as Hardy-Weinberg equilibrium, do not apply to the hemizygous alleles of chromosome Y. Lastly, Y chromosome reference panels for imputation are not widely available for GWAS (Anderson et al. 2019). Therefore, autosomes and sex chromosomes are typically analyzed separately.

Chromosome Y is unique in that it is passed exclusively from father to son and only recombines in pseudoautosomal regions (PARs) that make up ~5% of its DNA. As a result, ~95% of chromosome Y is identical between father and son, making it easy to identify ancestry and assign entire Y chromosomes to haplogroups. These haplogroups, sometimes referred to as clades, are defined by unique genetic markers and have been used to identify associations with disease. For example, specific Y chromosome haplogroups have been associated with AIDS progression, coronary artery disease, and infertility in males (Sezgin et al. 2009; Charchar et al. 2012; Ran et al. 2013; Eales et al. 2019). Therefore, the usage of Y chromosome haplogroups to identify associations with PD is a valid strategy when using large sample sizes. Here we take advantage of the structure of chromosome Y to identify haplogroups, variants and gene expression patterns potentially associated with PD risk in males using multiple large cohorts.

## 2. Methods

### 2.1 AMP-PD Data

We used Y chromosome whole genome sequencing (WGS) data from the Accelerating Medicines Partnership - Parkinson’s disease initiative (AMP-PD, https://amp-pd.org/) which includes data from multiple cohorts (Iwaki, Leonard, et al. 2021). This data contained a total of 184,317 Y chromosome variants and 9,887 samples, including 4,146 controls, 2,844 PD cases, 2,628 Lewy body dementia (LBD) cases, and 269 samples with a neurological disorder other than PD or LBD. Plink (v1.9) was used to filter this data (Chang et al. 2015). Female samples were removed, leaving 5,470 male samples totalling 1,895 controls, 1,751 PD cases, 1,668 LBD cases and 156 samples with another neurological disorder. Of these 5,470 males samples, 5,352 had European ancestry, which were included for further analyses. Heterozygous Y chromosome variants were set to missing and completely removed from all samples if they were heterozygous in over 10% of the male samples, bringing the variant count down to 183,109. To match the hg19 reference genome used in the haplogroup calling tools we used the UCSC liftover web tool (https://genome.ucsc.edu/cgi-bin/hgLiftOver) to lift the hg38 AMP-PD data to the hg19 human genome reference build. We removed variants that failed to convert (41,081 variants) and variants that converted to a different chromosome (345 variants) leaving us with a total of 141,683 variants. Non-European samples, samples with LDB, PD genetic carriers of known disease-causing variants identified from biased recruitment and samples with a neurological disorder other than PD were removed from the AMP-PD PD case/control dataset, leaving a total of 3,130 samples, including 1,466 cases and 1,664 controls.

### 2.2 UK Biobank Data

Y chromosome un-imputed genotype data was obtained from the UK Biobank (UKBB) (Bycroft et al. 2018). This contained 488,377 samples and 691 Y chromosome variants. PD phenotypes were based on data field 131023 (codes 40, 41, 50, 51, 20, 30 or 31) and proxy PD phenotypes (individuals without PD but with an affected father with PD) were based on data field 20107 (code 11). Female (based on genetic sex code 22001) and non-European samples were removed using Plink (v1.9) leaving 188,192 samples. Of the remaining male European samples there were 182,517 controls, 1,892 PD cases, and 3,783 proxy cases. There were about twice as many proxy samples as cases in the UKBB data, so one third of the total 182,517 controls were randomly selected for the UKBB PD case/control dataset, leaving 1,892 PD cases and 60,839 controls. The remaining two thirds of the controls were included in the UKBB proxy/control dataset, including 3,783 PD proxies and 121,678 controls.

### 2.3 NeuroX Data

NeuroX data was downloaded from dbGAP (phs000918.v1.p1) (Nalls et al. 2015). This dataset included 7,791 cases and 9,036 controls. The NeuroX data contained 139 Y chromosome genotyped variants. After standard sample level quality control was performed, including ancestry and relatedness, as reported elsewhere (Nalls et al. 2019), 3,491 cases and 3,232 controls were left, all of which had European ancestry.

### 2.4 Haplogroup Calling Tools

After sample and variant level quality control, AMP-PD, UKBB and NeuroX data were used to assign haplogroups to each sample. Three haplogroup calling tools were applied for comparison. Plink binary files were used as input for the Single Nucleotide Assignment of Phylogenetic Parameters on the Y chromosome (Snappy) tool (Severson et al. 2018). Plink binary files were converted to vcf format for the Yhaplo and Y-LineageTracker tools (Poznik 2016; Chen et al. 2021). Using output from these tools, each sample was assigned a major haplogroup by extracting the first character from each full haplogroup name (ex: R1a1a1b to R, J2a1b1 to J, etc) to follow grouping used by the International Society of Genetic Genealogy Y-DNA Haplogroup Tree (“International Society of Genetic Genealogy” 2020).

To verify the accuracy of the haplogroups assigned to each AMP-PD sample, we compared haplogroups of all related samples. Plink2 was used to filter for a minimum allele count of 2 and Hardy-Weinberg equilibrium of 0.0001 before calculating relatedness coefficients using the Kinship-based Inference for Genome-wide association studies (KING) tool (Manichaikul et al. 2010; Chang et al. 2015). We identified 129 pairs of related male samples after filtering for a minimum relatedness coefficient of 0.088. Of these, 124 pairs were assigned the same major haplogroup, and five pairs had different major haplogroups (Sup. Table 1). The relatedness coefficients for these five samples were between 0.0884 and 0.177, suggesting these were second degree relatives who inherited their Y chromosome from different fathers and are related through female relatives.

### 2.5 Statistical Analyses

Plink was used to calculate Y-specific principal components for all AMP-PD samples, including genetic PD carriers, using only Y chromosome variants to determine if samples cluster by major haplogroup (Figure 3). AMP-PD and NeuroX autosomal principal components were calculated with Plink and calculated with flashpca/2.0 for UKBB data (Abraham, Qiu, and Inouye 2017) using autosomal variants from all four datasets, including AMP-PD PD case/control, UKBB PD case/control, UKBB PD proxy/control and NeuroX case/control datasets. The Python statsmodels/0.12.1 package was used to perform logistic regression on four datasets to determine if haplogroup could predict disease status in males. The first five autosomal principal components for each dataset and age (age at baseline for AMP-PD, age at recruitment for UKBB and age at onset and age at recruitment for NeuroX cases and controls respectively) were included as covariates in this analysis.

The results of the logistic regression for each major haplogroup were meta-analyzed using inverse variance weighting under a fixed effect model with R/3.6 and the metafor/3.0-2 package (Viechtbauer 2010). The AMP-PD PD case/control, UKBB PD case/control, UKBB PD proxy/control and NeuroX datasets were included in this meta analysis. Data for a major haplogroup was excluded if there were fewer than fifty samples with the major haplogroup in the dataset.

Logistic regression was also performed for each full haplogroup that was in more than fifty samples in any of the four datasets. Results from the AMP-PD PD case/control, UKBB PD case/control, UKBB PD proxy/control and NeuroX datasets were meta-analyzed using inverse variance weighting under a fixed effect model with R/3.6 and the metafor/3.0-2 package (Viechtbauer 2010). Full haplogroups were only included in this meta analysis if they were present in more than fifty samples in at least two of the datasets, where the UKBB PD case/control and UKBB PD proxy/control datasets counted as one. This meta analysis included 17 unique haplogroups that were present in more than fifty samples. This was performed for each of the three haplogroup calling tools because haplogroup reference data differed between the tools. Haplogroup reference data from each tool was used to identify variants associated with each haplogroup. Variant names were merged with the comprehensive list of Y chromosome haplogroup variants provided by the International Society of Genetic Genealogy to identify the most up-to-date haplogroup name for each variant and account for differences in haplogroup names between the haplogroup calling tools. These were annotated with ANNOVAR (Wang, Li, and Hakonarson 2010) to identify variant function.

Single variant testing was performed for each dataset independently using logistic regression. A minor allele frequency filter of 0.05 was applied to each dataset before performing logistic regression with covariates of age, one-hot encoded major haplogroup and the first five autosomal principal components. One-hot encoded major haplogroups were only included as covariates if the major haplogroup was present in fifty or more samples in the dataset to reduce instances of collinearity. Few genetic markers were available for both UKBB (691 variants) and NeuroX (139 variants), but AMP-PD WGS data had full chromosome Y sequencing data available (141,683 variants). Association results across the AMP-PD PD case/control, NeuroX case/control, UKBB case/control and UKBB proxy/control datasets were combined and filtered to include only variants that were present in at least two of these datasets, bringing the variant count from 3320 down to 31. These 31 variants were meta-analyzed using inverse variance weighting under a fixed effect model with METAL (Willer, Li, and Abecasis 2010) and annotated with ANNOVAR. The AMP-PD PD case/control dataset was analyzed on its own since it was the only dataset with full coverage of the genome. This separate analysis utilized data prior to lifting over to hg19 to include more variants (183,109 variants). Insertions, deletions and multiallelic variants were removed prior to performing this analysis due to the high sequencing false positive rate of these variant types, leaving 994 variants.

### 2.6 RNA sequencing data

RNASeq data from the North American Brain Expression Consortium (NABEC) was used to quantify gene expression in the frontal cortex in 171 male samples, consisting entirely of population controls (Gibbs et al. 2010). Gene expression was quantified twice to determine if the exclusion of Y chromosome PARs, the only regions of the Y chromosome that recombine with the X chromosome, alters expression levels. To do so we used two different reference genomes, a version of the GENCODE release 38 hg38 reference genome with Y PARs masked, and the default GENCODE release 38 hg38 reference genome, which includes Y PARs by default (Frankish et al. 2021).

XYalign was used to mask Y PARs in one of the reference genome files (Webster et al. 2019). The genomeGenerate runMode from STAR was then used to generate a genome file containing masked Y PARs using the output from the XYalign tool and an annotated transcript file from GENCODE (Dobin et al. 2013). We followed the same parameters used by AMP-PD to generate reference genomes in anticipation of comparing expression between NABEC and AMP-PD samples. Similar steps were taken to generate a reference genome file including Y PARs, the only difference being the default GENCODE files including Y PARs as input. NABEC male samples were mapped to both the Y PAR masked reference genome and the default reference genome using the alignReads runMode in STAR. The featureCounts tool from the Subread package was used to count mapped reads for genes in all samples (Liao, Smyth, and Shi 2019), following the same parameters used in the AMP-PD RNASeq pipelines. The featureCount data was combined into two expression matrices, one containing counts for samples mapped to the reference genome with Y PARs masked and one containing counts for the samples mapped to the default reference genome. Each count matrix included a total of 171 samples and 60,708 genes, 566 of which were located on chromosome Y. The edgeR R package was used to filter out genes with low expression, leaving a total of 22,049 genes, and identify genes differentially expressed between the Y PAR masked and unmasked data (Robinson, McCarthy, and Smyth 2010). This analysis was followed up by thresholded testing using the glmTreat function, which repeated the test for differential expression, but relative to a minimum log fold change of two, instead of the default log fold change of zero. This thresholded testing was applied to all other differential expression analyses as well.

Blood based RNASeq data was obtained from AMP-PD to compare Y chromosome gene expression in blood with the previously quantified brain expression data. Expression data quantified with featureCounts was available at baseline for 563 Y chromosome genes and 1,020 AMP-PD samples after removing PD genetic carriers and non-European samples. This was combined with our requantified NABEC frontal cortex expression data generated using a reference genome including Y PARs. A total of 553 genes were common between the two datasets, including 1,020 blood samples and 171 brain samples. EdgeR was used to filter out genes with low expression, leaving a total of 275 genes, and to identify genes differentially expressed in brain and blood tissues in all samples while adjusting for major haplogroup.

EdgeR was used to identify Y chromosome genes differentially expressed between PD cases and controls in AMP-PD samples while adjusting for both sample age and major haplogroup. Genes with low expression were removed prior to analysis, leaving 278 genes for the 1,020 AMP-PD samples, including 722 cases and 298 controls.

A similar analysis was done to identify genes differentially expressed between major haplogroups in AMP-PD samples while adjusting for case/control status. Genes with low expression were filtered out prior to performing this analysis, leaving a total of 240 genes for the 1,020 AMP-PD samples. This was only performed for major haplogroups present in at least 40 samples, leaving major haplogroups E, G, I, J, and R. Gene expression for samples with each of these major haplogroups was compared with expression for all samples with a different major haplogroup. Gene counts were plotted for Y chromosome genes of interest along with case control status and sample major haplogroup obtained from Yhaplo to visualize differences in gene expression. This was repeated with the NABEC frontal cortex expression, excluding case/control status because all samples were controls (Figure 4).

### 2.7 Data and Code availability

All AMP-PD (https://amp-pd.org/) and UKBB (https://www.ukbiobank.ac.uk/) data is available via application on their websites and the NeuroX data is available via dbGap at phs000918.v1.p1. NABEC data is available via dbGap at phs001354.v1.p1. All code is available on the GitHub page: https://github.com/neurogenetics/chrY_haplogroups_PD

## 3. Results

### 3.1 Identification of chromosome Y haplogroups

In total we included 6,849 PD cases, 3,783 proxy cases and 187,413 controls derived from AMP-PD, UKBB and NeuroX cohorts. We called haplogroups for all included samples using three different tools: Snappy, Yhaplo, and Y-LineageTracker (Figure 1). Full haplogroup frequencies, major haplogroup frequencies and the number of unique haplogroups identified by each tool using each cohort were recorded to compare across tools (Sup. Tables 2-7, 10, Figure 2). Overall, good concordance was observed.

**Figure 1.**
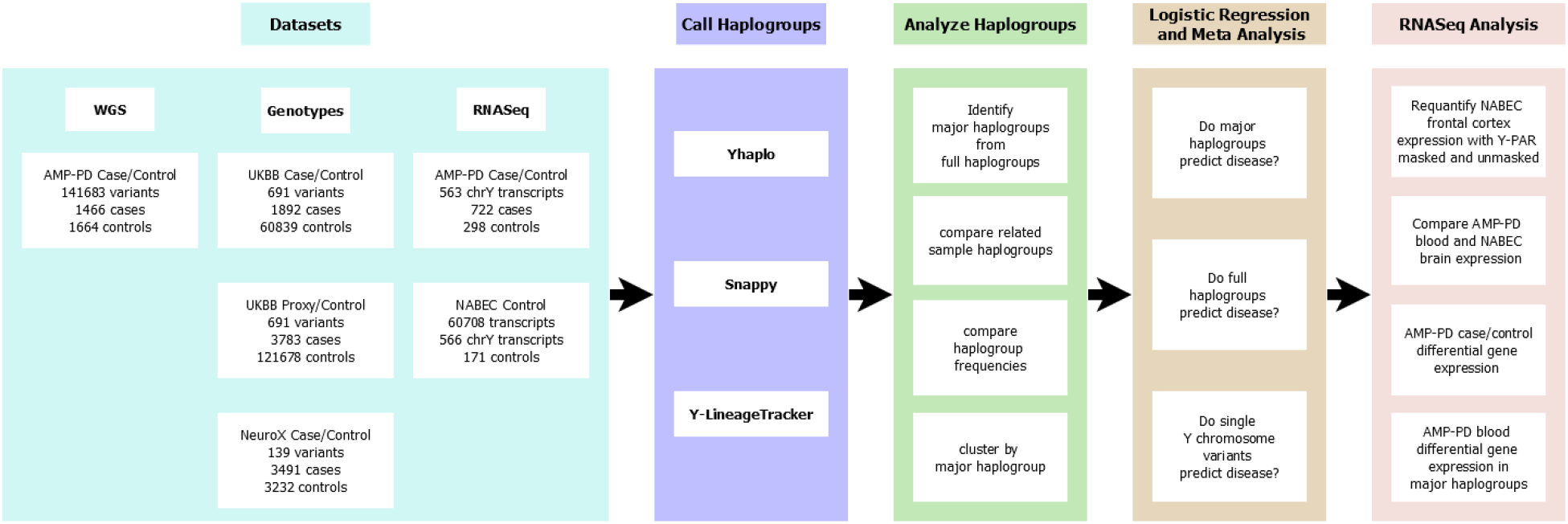
Analysis flow-chart of included data and performed analyses.

**Figure 2.**
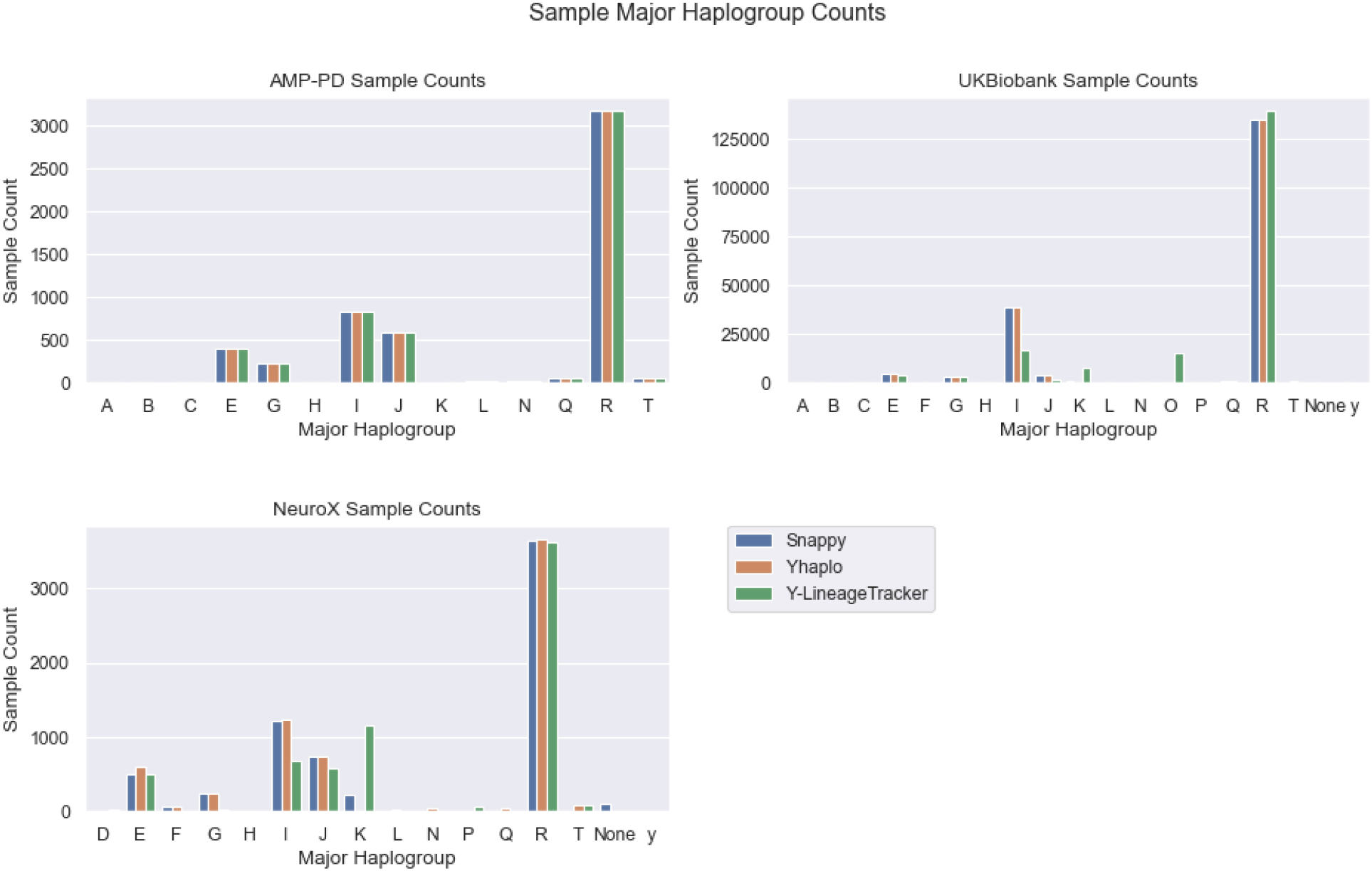
Comparison of major haplogroup counts in Y chromosome haplogroup calling tools. Major haplogroup counts using AMP-PD whole genome sequencing data (top left), major haplogroup counts using UKBiobank genotype data (top right), and major haplogroup counts using NeuroX genotype data (bottom left).

Y chromosome major haplogroup frequencies were obtained from six different studies to compare our frequencies with those of European populations in specific countries (Sup. Table 15). These included Belgium, the Netherlands (Larmuseau et al. 2011), Poland (Grochowalski et al. 2020), Spain, Portugal (Adams et al. 2008), Belarus (Kharkov et al. 2005), the United Kingdom (Bowden et al. 2008), Algeria, Egypt, and Italy (Bekada et al. 2013). Major haplogroup frequencies identified in European ancestry samples using Yhaplo for the AMP-PD, UKBiobank and NeuroX cohorts were similar to those of other European countries.

To visualize the differences between haplogroups, Y-specific principal components were calculated for AMP-PD samples using Y chromosome data. The first two principal components were plotted with the major haplogroups obtained from the Snappy and Yhaplo tools which both assigned identical major haplogroups to all AMP-PD samples. Since haplogroups A and B are the oldest (Karmin et al. 2015; Y Chromosome Consortium 2002), initial clustering focused on these two major haplogroups (Figure 3A). Removal of these haplogroups shows clear clustering based on identified major haplogroup (Figure 3B).

**Figure 3.**
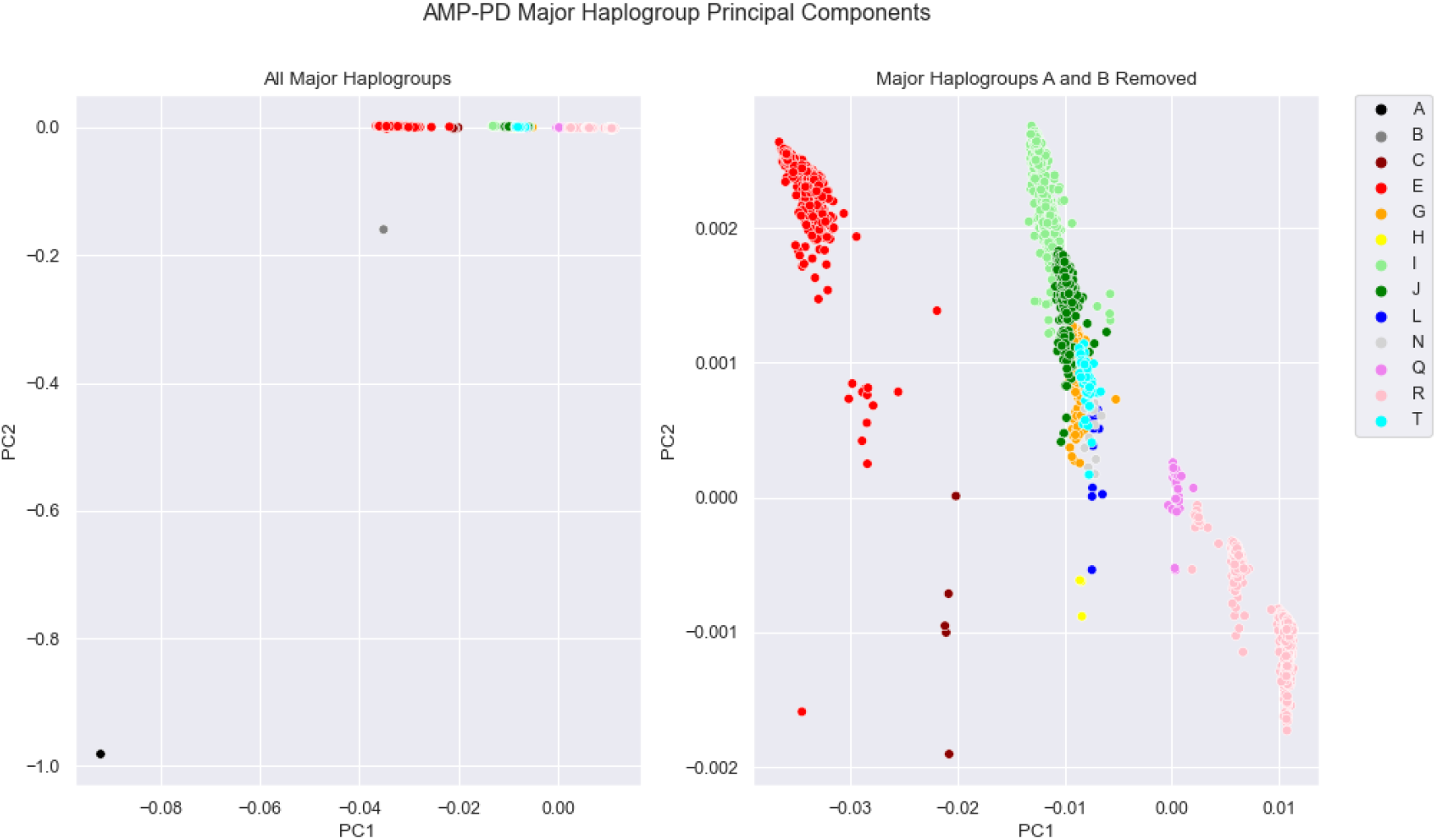
Y Chromosome Genetic Principal Components with Y Chromosome Major Haplogroups. The first two principal components are plotted with all major haplogroups (left) and with major haplogroups A and B removed (right).

### 3.2 Case-control chromosome Y associations

Logistic regression was performed on each dataset for each major haplogroup obtained from Yhaplo to determine if a major haplogroup can predict disease status (Sup. Table 8). These results were combined to perform a meta analysis for each major haplogroup (Sup. Table 9). The meta analysis only included datasets with at least fifty samples with the major haplogroup. No major haplogroups had significant association (p-value < 0.05) with PD after performing meta analysis.

Logistic regression was conducted on each dataset for each full haplogroup obtained from all three haplogroup calling tools to identify variants making up haplogroups that predict disease status. Logistic regression was only performed on haplogroups that were in fifty or more samples in the dataset (Sup. Table 11). These results were combined to perform a meta analysis for each full haplogroup present in at least two of the cohorts. Of these, one haplogroup (R1b1a2a1a) was significant after multiple test correction (p-value < 0.05/17=0.002) (Sup. Table 12). Variants representing this haplogroup were obtained from each tool’s haplogroup reference data and annotated with ANNOVAR (Wang, Li, and Hakonarson 2010) to determine variant function. All variants associated with the single significant full haplogroup, R1b1a2a1a, which was identified by the Yhaplo tool, were intergenic. R1b1a2a1a is also known as R-L11 and R-L151 and is predominantly found in western European countries. However, some inconsistencies were identified for this specific haplogroup between the methods used for Y chromosome haplogroup calling (Sup. Table 11) so caution should be taken into consideration when interpreting these results.

### 3.3 Single variant testing

Plink logistic regression was performed to identify Y chromosome variants associated with PD while adjusting for covariates of age, one-hot encoded major haplogroup and the first five autosomal principal components. METAL was used to meta-analyze the AMP-PD PD case control, UKBB PD case control, UKBB PD proxy control, and NeuroX PD case control datasets together. However given the lack of chromosome Y coverage in the UKBB and NeuroX cohorts, very few variants could be meta-analyzed (Sup. Table 13). In total we meta-analyzed 31 variants and none passed correction for multiple testing. A separate analysis of the AMP-PD data identified a total of 11 Y chromosome variants with p-value < 0.05, but none passed multiple test corrections (Sup. Table 14).

### 3.4 Gene expression assessments of different Y chromosome haplogroups

In addition to assessing genetic associations between chromosome Y and PD, we investigated 1) if the PARs affect gene quantifications of Y chromosome genes, 2) whether blood and brain gene expression of the Y chromosome are comparable and 3) whether gene-expression differences of the Y chromosome are associated with PD or Y chromosome haplogroup.

Previous studies have suggested that the removal of the Y chromosome, including Y PARs, from the reference genome improves mapping quality and alters downstream variant calling results across the X chromosomes of female samples (Webster et al. 2019). To determine if similar behavior exists on the Y chromosome of male samples, we measured differences in Y chromosome gene expression when masking Y PARs in male samples. To do so we requantified frontal cortex expression in NABEC samples using a Y PAR masked reference genome and a reference genome including Y PARs. EdgeR was used to compare the results of these two methods. Thresholded testing was applied using the glmTreat function to test for differential expression relative to a minimum log fold change of 2, instead of the default log fold change of zero. This resulted in 17 differentially expressed genes (Sup. Table 16). All of these genes were upregulated in samples mapped to the Y PAR masked reference genome and were located in PARs. Therefore, we concluded that the presence of Y PARs would not impact any additional analysis of Y chromosome gene expression.

Next we conducted a linear regression with edgeR to identify Y chromosome genes differentially expressed between brain and blood tissues to assess whether blood expression could be used as a proxy for brain gene expression. Using blood expression from the 1,020 AMP-PD samples and brain expression from the 171 NABEC samples, we identified a total of 207 genes upregulated in blood tissues, 51 genes upregulated in brain tissues and 17 non-significant genes. Thresholded testing relative to a log fold change of two was applied, resulting in a total of 160 genes upregulated in blood tissues, 25 genes upregulated in brain tissues, and 90 non-significant genes, suggesting blood is a poor proxy for brain tissue when quantifying gene expression (Sup. Table 17).

Finally, we assessed whether gene-expression differences of the Y chromosome are associated with PD or Y chromosome haplogroup. Genes differentially expressed in PD cases and controls were identified with edgeR using the blood derived AMP-PD data. One pseudogene, *KDM5DP1*, was significantly upregulated in cases, but did not remain significant after thresholded testing relative to a log fold change of 2 was applied (Sup. Table 18). Next, differences in Y chromosome gene expression between major haplogroups were identified in AMP-PD samples using edgeR. Only major haplogroups present in at least 40 samples were compared, leaving major haplogroups E, G, I, J and R (Sup. Figure 2). After filtering out genes with low expression we identified a total of 64 genes upregulated in individuals with major haplogroup G and eight genes upregulated in samples with major haplogroup J. Four of these 72 genes had a log fold change greater than two (Sup. Figure 3). None of the total 240 genes were differentially expressed in major haplogroup E, I and R samples when compared to samples with different haplogroups. However when we applied thresholded testing relative to a log fold change of 2, no genes were differentially expressed in any major haplogroup (Sup. Tables 19-23).

## 4. Discussion

Here we assessed whether Y chromosome variation contributes to PD using the largest available case-control publicly available PD datasets containing Y chromosome genotypes. We used three different algorithms to call Y haplogroups, which had high concordance rates for major haplogroups. Minor differences were identified in a small number of individuals, however the major haplogroup was typically consistent (Figure 2).

Major haplogroup R was the most common in all three of our cohorts (AMP-PD, UKBiobank, and NeuroX), present in at least half of all samples. Our comparison with other studies shows that the same is applicable for European countries such as Belgium, the Netherlands, the United Kingdom, Poland, Spain, Portugal and Belarus (Sup. Table 15). All included cohorts and these countries had the same major haplogroup with the second highest frequency (named I), with the exception of Spain and Portugal. It should be noted that major haplogroup frequencies in our included cohorts were distinct from frequencies found in the Northern African countries of

Algeria and Egypt and in areas of southern Europe, including Italy. Samples included in our study were obtained from European ancestry populations, so these comparisons support the accuracy and reliability of the Snappy, Yhaplo and Y-LineageTracker haplogroup calling tools for determining major haplogroups.

However, the same cannot necessarily be stated for the reliability of these tools when assigning full Y chromosome haplogroups. Of the three tools, Y-LineageTracker identified the most unique full haplogroups in the AMP-PD cohort (including NABEC samples) and Yhaplo identified the most unique full haplogroups in the UKBB and NeuroX cohorts (Sup. Table 10). This contrast is likely due to differences between algorithms used by the tools, differences in the curated lists of Y chromosome variants and haplogroups used by each tool, or the differences between chromosome Y variant coverage across included cohorts. Variants in these lists were counted and compared with the AMP-PD, UKBB, and NeuroX cohorts to further investigate the potential of these tools (Sup. Figure 1). Of the three tools, Y-LineageTracker included the largest number of Y chromosome variants used to nominate a haplogroup. Compared to variants used by the other tools, this list of variants had the most overlap (9,135 variants) with the variants present in the AMP-PD cohort, the largest cohort of the three (141,683 variants). However, only a small percentage (less than 20%) of the variants explored through each tool were present in each of the three cohorts. Therefore, it is possible that precise Y chromosome haplogroups were incorrectly assigned for some samples. However, the similarity of major haplogroup frequencies with past studies suggests these tools remain reliable when identifying major haplogroups.

Interestingly, Yhaplo had the largest percentage of variants in its reference file included in all three cohorts (~19% for AMP-PD, ~1.7% for UKBB, ~0.5% for NeuroX). Therefore, Yhaplo was used to determine major haplogroups in our analyses. The large difference in percentages suggests that WGS data, such as the AMP-PD data, is a much better fit for our analyses than genotype data, such as the UKBB and NeuroX data. The performance of these tools will likely increase as methods to identify Y chromosome variants improve, such as the development of Y chromosome imputation panels, allowing for a better understanding of the relationship between Y chromosome haplogroups and PD.

Meta-analysis of the logistic regression per haplogroup identified no major haplogroups associated with PD (Sup. Table 9). A meta-analysis focusing on the more specific full haplogroups resulted in one haplogroup associated with PD after multiple test correction, however discordant results were identified between Y chromosome calling tools used. Our approach suggests that caution needs to be taken into consideration with these results and additional replication is needed (Sup. Table 12). No Y chromosome variants were found to be associated with disease after multiple test correction in an analysis of AMP-PD data or in a meta-analysis of all datasets (Sup. Tables 13, 14). Therefore, our analyses identified neither Y chromosome haplogroups nor Y chromosome variants significantly associated with PD.

While Y chromosome gene expression was overall low in both brain and blood tissues, we identified clear differences in Y chromosome gene expression between these tissues both before and after applying thresholded testing with a log fold change filter to the edgeR results (Sup. Table 17). This is in line with previous studies that have shown Y chromosome genes to be most highly expressed in sex specific tissues (Godfrey et al. 2020).

A large proportion of genes were removed from the datasets used in each differential expression analysis when genes with zero expression in all samples were removed and when the filterByExpr edgeR function was used. After applying these filters, a total of 22,049 of the 60,708 (including autosomal genes) NABEC genes remained for the Y PAR analysis, 275 of the 553 genes remained for the NABEC brain vs AMP-PD blood analysis, 278 of the 563 AMP-PD genes remained for the PD case control analysis, and 240 of the 563 AMP-PD genes remained for the major haplogroup analysis.

Comparison of gene expression between samples mapped to a reference genome without Y PARs and samples mapped to a reference genome with Y PARs identified a total of 17 genes significantly upregulated in the samples without Y PARs. All of these genes were located in PAR regions on the X chromosome. This was expected, because Y PARs were not present in the reference genome used for these samples. Gene expression was not significantly different for genes in any other areas of the genome. This suggests that masking PARs when quantifying gene expression likely does not significantly alter expression levels of genes outside of PARs when using brain derived datasets.

Differences in gene expression between major haplogroups were identified in major haplogroups G and J (Sup. Tables 20 and 22). Genes with high log fold change values displayed clear differences in expression levels between samples (Sup. Figure 3). However, none of these genes remained significant after thresholded testing was applied, suggesting gene expression patterns are not specific to Y chromosome haplogroups. There was a similar lack of differentially expressed genes in the PD case/control analysis, suggesting Y chromosome gene expression patterns are not specific to disease status (Sup. Table 18).

To date, chromosome Y PD studies have focused on several genes, including *SRY*. In vitro studies suggest upregulation of *SRY* protects against 6-hydroxydopamine induced PD in male dopaminergic neurons (Czech et al. 2014). An in vivo study has demonstrated that inhibition of *SRY* diminishes dopaminergic cell damage in 6-hydroxydopamine and rotenone induced PD rat models (Lee et al. 2019). A study of PD in Asian populations has found no significant association between *SRY* variants and PD risk (Pan et al. 2021). In concordance with this study, we found no significant *SRY* variants to be associated with PD and expression patterns of *SRY* in brain and blood tissue was overall low, suggesting this gene does not play a major role in PD (Figure 4). Mouse studies have shown that *SRY* expression is precisely regulated during embryonic development, going from high to low expression in a few days, to properly trigger male development (Larney, Bailey, and Koopman 2014). This suggests an assessment of *SRY* expression in the brain of older individuals, like in our study, may be of limited use. Additionally, *SRY* indirectly leads to the production of testosterone, a steroid found to be significantly lowered in human PD cases (Toczylowska et al. 2020). Findings such as these suggest *SRY*, or other genes involved in sex determination or hormone regulation, may indirectly influence male neurobiology, potentially explaining the increased prevalence of PD in males (Kopsida et al. 2009).

**Figure 4.**
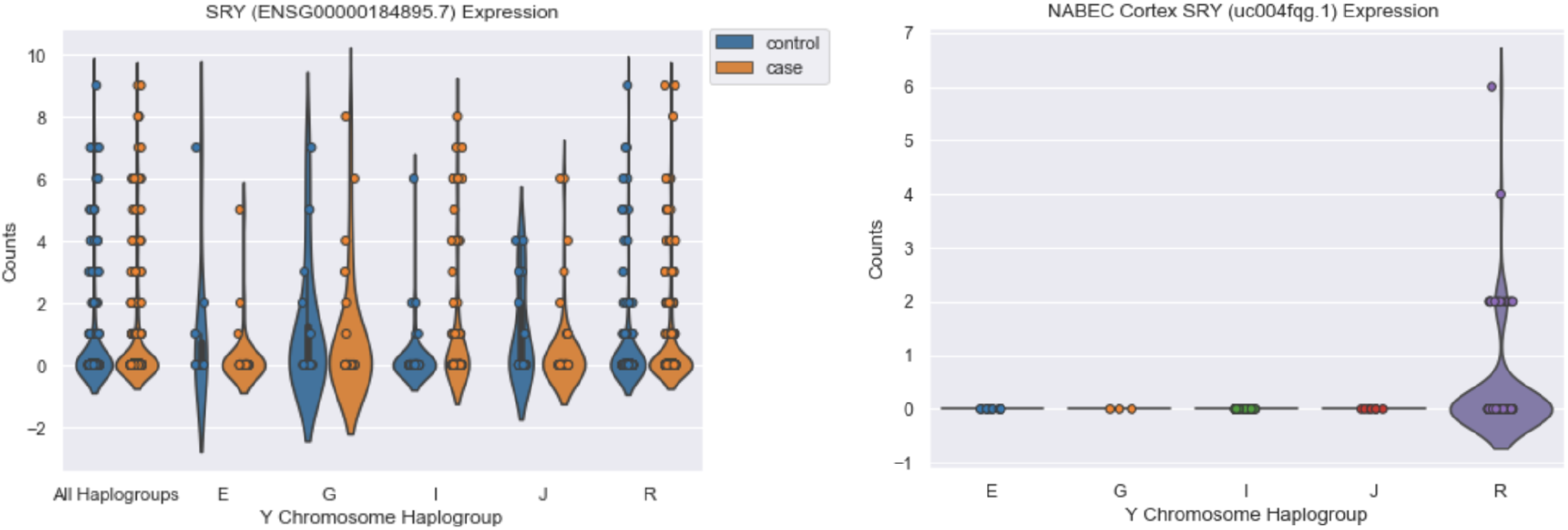
*SRY* expression in blood from AMP-PD samples and frontal cortex from NABEC samples. *SRY* counts obtained from featureCounts grouped by case/control status and major haplogroup in blood tissues using AMP-PD data (left) and in brain tissues using NABEC frontal cortex data (right).

Overall, our data suggests that genetic variation on chromosome Y does not have a major direct effect on risk for PD. However, it is possible that regulatory Y chromosome variants indirectly affect PD risk. Future research will need to reassess this scientific question once additional WGS data and more genetically diverse data is available.

## Data Availability

https://www.ukbiobank.ac.uk/

https://github.com/neurogenetics/chrY_haplogroups_PD

## Acknowledgements and Funding

We would like to thank all of the subjects who donated their time and biological samples to be part of this study. This work was supported in part by the Intramural Research Programs of the National Institute on Aging (NIA) part of the National Institutes of Health, Department of Health and Human Services; project numbers 1ZIA-NS003154, Z01-AG000949-02, ZO1 AG000535 and Z01-ES101986. We are grateful to members of the North American Brain Expression Consortium (NABEC) for contributing DNA samples. This research has been conducted using the UK Biobank Resource under Application Number 33601.

Data used in the preparation of this article were obtained from the Accelerating Medicine Partnership® (AMP®) Parkinson’s Disease (AMP PD) Knowledge Platform. For up-to-date information on the study, visit https://www.amp-pd.org. The AMP® PD program is a public-private partnership managed by the Foundation for the National Institutes of Health and funded by the National Institute of Neurological Disorders and Stroke (NINDS) in partnership with the Aligning Science Across Parkinson’s (ASAP) initiative; Celgene Corporation, a subsidiary of Bristol-Myers Squibb Company; GlaxoSmithKline plc (GSK); The Michael J. Fox Foundation for Parkinson’s Research; Pfizer Inc.; Sanofi US Services Inc.; and Verily Life Sciences. ACCELERATING MEDICINES PARTNERSHIP and AMP are registered service marks of the U.S. Department of Health and Human Services. Clinical data and biosamples used in preparation of this article were obtained from the Michael J. Fox Foundation (MJFF) and National Institutes of Neurological Disorders and Stroke (NINDS) BioFIND study, Harvard Biomarkers Study (HBS), the NIA International Lewy Body Dementia Genetics Consortium Genome Sequencing in Lewy body dementia case-control cohort (LBD), the MJFF LRRK2 Cohort Consortium (LCC), the NINDS Parkinson’s disease Biomarkers Program (PDBP), MJFF Parkinson’s Progression Marker Initiative (PPMI), and the NINDS Study of Isradipine as a Disease Modifying Agent in Subjects With Early Parkinson Disease, Phase 3 (STEADY-PD3). BioFIND is sponsored by The Michael J. Fox Foundation for Parkinson’s Research (MJFF) with support from the National Institute for Neurological Disorders and Stroke (NINDS). The BioFIND Investigators have not participated in reviewing the data analysis or content of the manuscript. For up-to-date information on the study, visit michaeljfox.org/news/biofind. Genome sequence data for the Lewy body dementia case-control cohort were generated at the Intramural Research Program of the U.S. National Institutes of Health. The study was supported in part by the National Institute on Aging (program #: 1ZIAAG000935) and the National Institute of Neurological Disorders and Stroke (program #: 1ZIANS003154). The Harvard NeuroDiscovery Biomarker Study (HBS) is a collaboration of HBS investigators [full list of HBS investigator found at https://www.bwhparkinsoncenter.org/biobank/ and funded through philanthropy and NIH and Non-NIH funding sources. The HBS Investigators have not participated in reviewing the data analysis or content of the manuscript. The LRRK2 Cohort Consortium is coordinated and funded by The Michael J. Fox Foundation for Parkinson’s Research. Data used in preparation of this article were obtained from the MJFF-sponsored LRRK2 Cohort Consortium (LCC). The LCC Investigators have not participated in reviewing the data analysis or content of the manuscript. For up-to-date information on the study, visit https://www.michaeljfox.org/biospecimens. PPMI – a public-private partnership – is funded by the Michael J. Fox Foundation for Parkinson’s Research and funding partners, including [list the full names of all of the PPMI funding partners found at www.ppmi-info.org/fundingpartners. The PPMI Investigators have not participated in reviewing the data analysis or content of the manuscript. For up-to-date information on the study, visit www.ppmi-info.org. Parkinson’s Disease Biomarker Program (PDBP) consortium is supported by the National Institute of Neurological Disorders and Stroke (NINDS) at the National Institutes of Health. A full list of PDBP investigators can be found at https://pdbp.ninds.nih.gov/policy. The PDBP Investigators have not participated in reviewing the data analysis or content of the manuscript. The Study of Isradipine as a Disease Modifying Agent in Subjects With Early Parkinson Disease, Phase 3 (STEADY-PD3) is funded by the National Institute of Neurological Disorders and Stroke (NINDS) at the National Institutes of Health with support from the Michael J. Fox Foundation and the Parkinson Study Group. For additional study information, visit https://clinicaltrials.gov/ct2/show/study/NCT02168842. The STEADY-PD3 investigators have not participated in reviewing the data analysis or content of the manuscript. The Study of Urate Elevation in Parkinson’s Disease, Phase 3 (SURE-PD3) is funded by the National Institute of Neurological Disorders and Stroke (NINDS) at the National Institutes of Health with support from the Michael J. Fox Foundation and the Parkinson Study Group. For additional study information, visit https://clinicaltrials.gov/ct2/show/NCT02642393. The SURE-PD3 investigators have not participated in reviewing the data analysis or content of the manuscript.” “The LRRK2 Cohort Consortium (LCC) was created to assemble and study groups of people with and without Parkinson’s disease who carry mutations in the LRRK2 gene. The LRRK2 Cohort Consortium is coordinated and funded by The Michael J. Fox Foundation for Parkinson’s Research. The investigators within the LCC contributed to the design and implementation of the LCC and/or provided data and/or collected biospecimens, but did not necessarily participate in the analysis or writing of this report. The full list of LCC investigators can be found at www.michaeljfox.org/lccinvestigators.

## COI

M.A.N. ‘s participation in this project was part of a competitive contract awarded to Data Tecnica International LLC by the National Institutes of Health to support open science research, he also currently serves on the scientific advisory board for Clover Therapeutics and is an advisor to Neuron23 Inc.

## Notes

### Funding Statement

This study was funded by the National Institutes on Aging.

### Author Declarations

All AMP-PD (https://amp-pd.org/) and UKBB (https://www.ukbiobank.ac.uk/) data is available via application on their websites and the NeuroX data is available via dbGap at phs000918.v1.p1. NABEC data is available via dbGap at phs001354.v1.p1.

