## Supplementary Figures for "Analysis of Y Chromosome Haplogroups in Parkinson’s Disease"

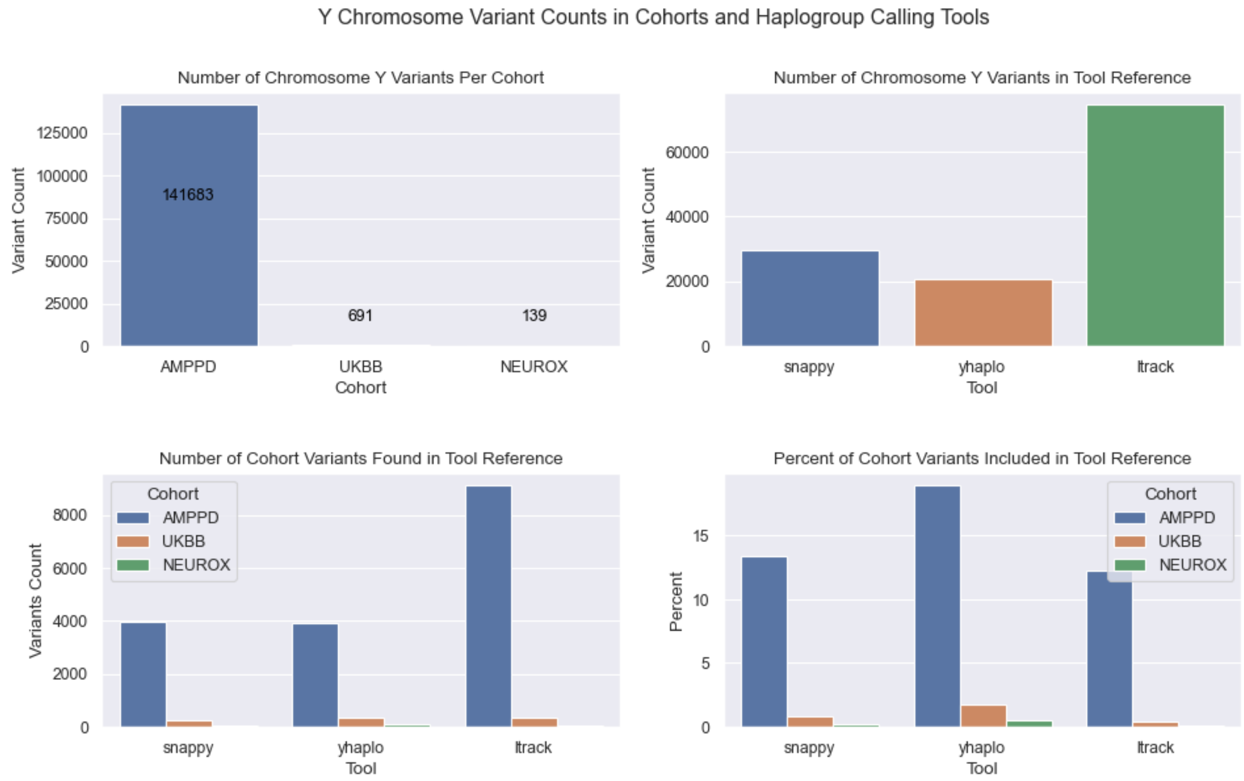


**Supplementary Figure 1. Comparison of Y chromosome variant counts in cohorts and haplogroup calling tools.** We compared the number of Y chromosome variants available in each cohort's dataset (top left). The number of Y chromosome variants included in each tool's reference files used to assign haplogroups were compared (top right). The number of Y chromosome variants found in each tool's reference file for each cohort were compared to each other (bottom left). The number of Y chromosome variants found in each tool's reference file divided by the number of Y chromosome variants in each tool's reference (bottom right).

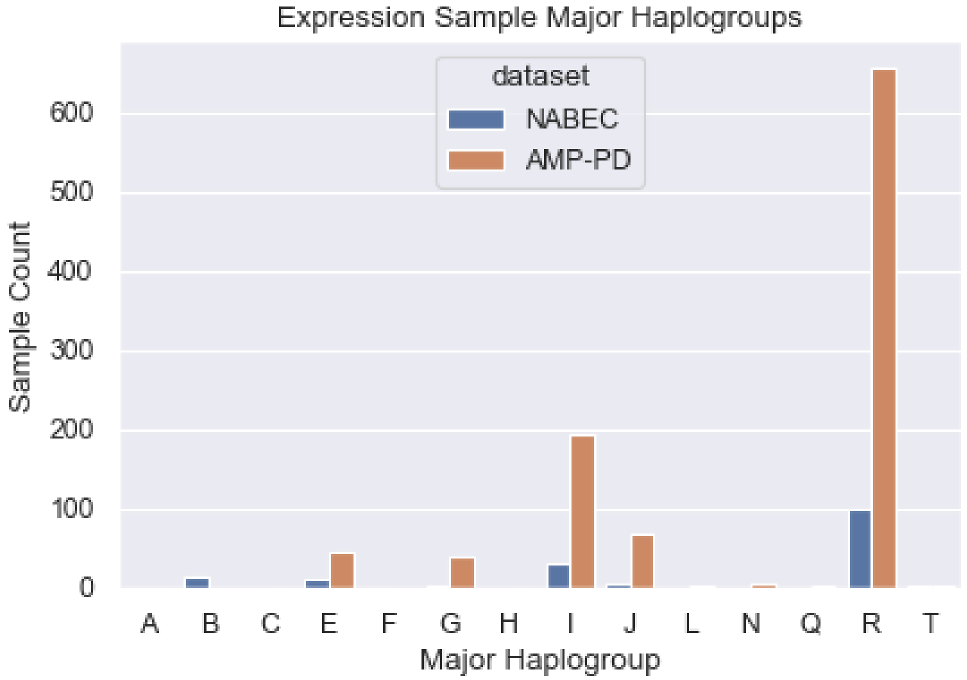


**Supplementary Figure 2. Sample major haplogroup counts in RNASeq data.** Number of samples with a specific major haplogroup in brain samples (NABEC) and blood samples (AMP-PD).

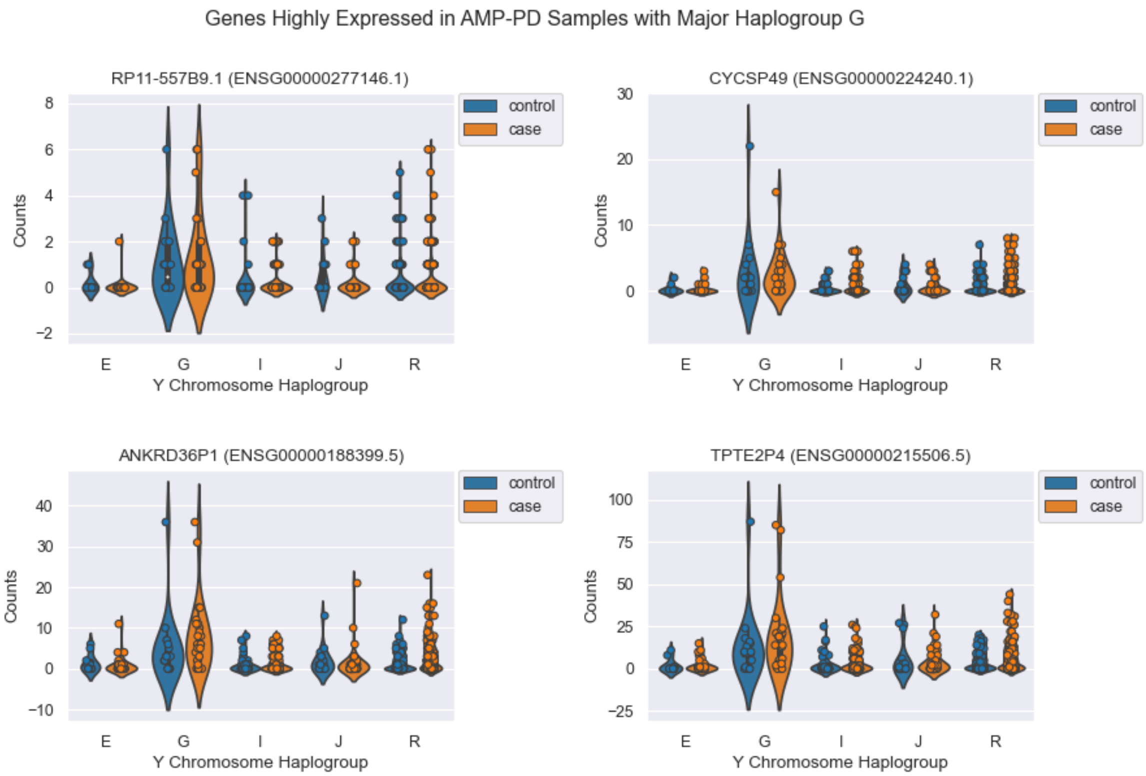


**Supplementary Figure 3. Gene counts for genes highly expressed in AMP-PD samples with major haplogroup G.** Gene counts obtained from AMP-PD featureCounts data grouped by sample case/control status and sample major haplogroup. Genes included passed multiple test correction and had a log fold change greater than two in edgeR results before thresholded testing was applied with the glmTreat function.
